# Stanford Screenomics: An Open-source Platform for Unobtrusive Multimodal Digital Trace Data Collection from Android Smartphones

**DOI:** 10.1101/2025.06.24.25329707

**Authors:** Ian Kim, Jack Boffa, Mujung Cho, David E. Conroy, Nathan Kline, Nick Haber, Thomas N. Robinson, Byron Reeves, Nilàm Ram

## Abstract

Smartphone-based digital trace data can offer powerful insights for identifying behavioral patterns and health risks. However, existing tools for comprehensive data collection lack scalability, customizability, transparency, and accessibility. To address these gaps, we developed an open-source platform that enables in-situ capture of multimodal digital traces from smartphones (e.g., moment-by-moment capture of screenshots, application usage logs, interaction histories, and phone sensor readings). The Stanford Screenomics Data Collection application allows researchers to tailor data types and quality, data transfer methods, and upload cadence. The Dashboard application supports real-time monitoring of participants’ data provision, identification of data issues, and automated reactive communications to participants. The platform’s back-end employs a NoSQL database for secure, and HIPAA-compliant storage. Using illustrative 24-hour digital trace data we demonstrate how the platform expands the range of possible digital phenotyping studies.

Date of Submission: [06] 2025

This article has not been published before and is not currently being considered for publication.

## INTRODUCTION

The digital context has emerged as a new social ecology, engaging large segments of the population for extended periods (Navarro & Tudge, 2023). However, understanding this digital ecology has been challenging, as traditional measures have been limited by relatively coarse time-based metrics. The recent rise in the collection of digital traces, defined as records of activity from online systems, has opened new avenues for researchers and clinicians to investigate a diverse collection of behaviors and activities that influence, are integrated with, and potentially support individuals’ health. The breadth of possible applications of digital traces is impressive and includes tracking of, for example, the development of interpersonal relationships (Allen et al., 2014; Pentland, 2014), patterns in online purchases (Audy Martínek et al., 2023; Kunst & Vatrapu, 2019), political orientations Chan et al., 2017; Kosinski et al., 2013), and education and learning (Du et al., 2023; Lämsä et al., 2025), adolescent mental health (Jensen et al., 2019; Orben & Przybylski, 2019), and behavioral health trends (Amir et al., 2019; Kim et al., 2023). Researchers in all these different domains are finding value in granular records of digital experiences that allow them to observe processes that have heretofore been invisible but are increasingly important for studying human behavior and supporting human health.

Digital trace data relevant to human behavior and health come from various sources, including individuals’ smartphones, wearable devices, and laptops. Among those, smartphone-based digital traces have become particularly useful due to the widespread adoption of smartphones. The ability to capture fine-grained, objective, and continuous data from smartphones about the owners’ physical, cognitive and social behaviors and the environments in which those behaviors manifest provides researchers and clinicians with a detailed, unobtrusive, and scalable window into a plethora of real-world human activities. Consequently, smartphone-based digital traces enable large-scale, cost-effective studies and population-level analyses, yielding unprecedented insights into human behavior and health (Eagle & Pentland, 2006; Gordon & Mendes, 2021; Lathia et al., 2017; K. Lee et al., 2023; Ohme et al., 2024). Notable examples include nationwide research on contact tracing, mobility trends, and behavioral shifts—such as changes in physical activity, screen time, and call and text logs—that can inform health policies and strategies (Kleinman & Merkel, 2020; Mejova, 2022).

Early on, smartphone-based digital traces focused on basic usage information, such as call logs and SMS metadata. Over time, the scope of digital traces expanded to include location and activity data from built-in GPS sensors, accelerometers and gyroscopes, as well as total screen time and app-specific usage. However, much of this “digital exhaust” only provides indirect insights into specific user behaviors rather than a comprehensive and direct view of users’ actual experiences and interactions with their fast-moving and idiosyncratic digital environment. Indeed, the literature on the impacts of indirect monitoring on smartphone use on health and behavior has been mixed (Twenge, 2017; Amez & Baert, 2020; Nam & Cha, 2024; Singh & Samah, 2018). The emerging realization is that the sensor metrics and usage logs, currently at the core of smartphone-based data collection, need to be supplemented with information about the actual content flowing through individuals’ smartphones. Many research questions remain unexplored simply because we lack information on what users are actually seeing and doing on their screens (Faust et al., 2024; Ohme et al., 2024; Yee et al., 2023).

A promising approach for bridging the gaps in knowledge is to also capture content-based data, such as smartphone screenshots and user-smartphone interaction logs of scrolling and swiping motions, alongside the other data streams (Ram et al., 2020; Reeves et al., 2021). This approach, known as Screenomics, advances traditional digital trace methods by offering a more direct, accurate, and comprehensive information about individuals’ digital lives and how they are connected to changes in health and well-being. A handful of recent studies illustrate the utility of these data traces for identifying patterns in production and consumption behavior (Cho et al., 2023), poverty-related differences in information flows (J. Lee et al., 2023), connections between adolescents and their parents (Sun et al., 2023), changes in frequency of interactions prior to a suicidal crisis (Jacobucci et al., 2024), idiosyncracy in content profiles (Brinberg et al., 2021), person-specific relations between patterns of smartphone use and mental health (Cerit et al., 2025), and developing personalized real-time digital context-dependent interventions (Ram et al., 2024). These studies exemplify the value of collecting granular information about smartphone experiences, some of which only last seconds, that may be critical to understanding the interplay between digital and real-life experiences.

Beyond these specific studies, comprehensive digital trace data create unique opportunities to advance and inform health research, interventions and policy. Field observations, laboratory studies, clinical trials, population health studies, and precision medicine approaches have often relied on self-report surveys or basic device logs to measure digital behaviors and exposures. These data streams do provide insight into some general aspects of digital life, but with limited resolution and without contextual information. Newly available comprehensive digital trace data captured from smartphones, including details of exposure to and engagement with particular virtual environments and content, online social interactions, activity spaces (e.g., particular applications), and human-computer interactions, can now be collected alongside and integrated with electronic health records, imaging, molecular and omics profiles, physiological biomarkers, wearable sensor outputs, and other biomedical data. The combined data extends the possibilities for a wide variety of research and practical applications, including the development of personalized and precisely-targeted interventions.

Figure 1 illustrates how comprehensive digital trace data, together with traditional health measures, contribute to and inform various health research and application domains. Comprehensive digital trace data serve as a form of continuous field observation, capturing behavioral and contextual data in real time in individuals’ natural environments. These data allow researchers to link moment-to-moment variations in daily life with health outcomes, detect early warning signals, and generate hypotheses and tests that were previously impossible to formulate or operationalize. For example, irregular sleep or disrupted sleep associated with evening social media content consumption may influence inflammatory markers or circadian gene expression. In clinical trials, randomly allocated interventions may be delivered in response to specific behavioral or environmental contexts and, to identify mechanisms of action, temporal patterns of hypothesized or exploratory activity and contextual mediators to help explain variability in adherence and physiological responses in response to behavioral, pharmacological, or surgical interventions. At the population level, combining these data with surveys or registries supports scalable monitoring, early detection of emerging risks, and timely assessment of behavioral dynamics.

**Figure 1.**
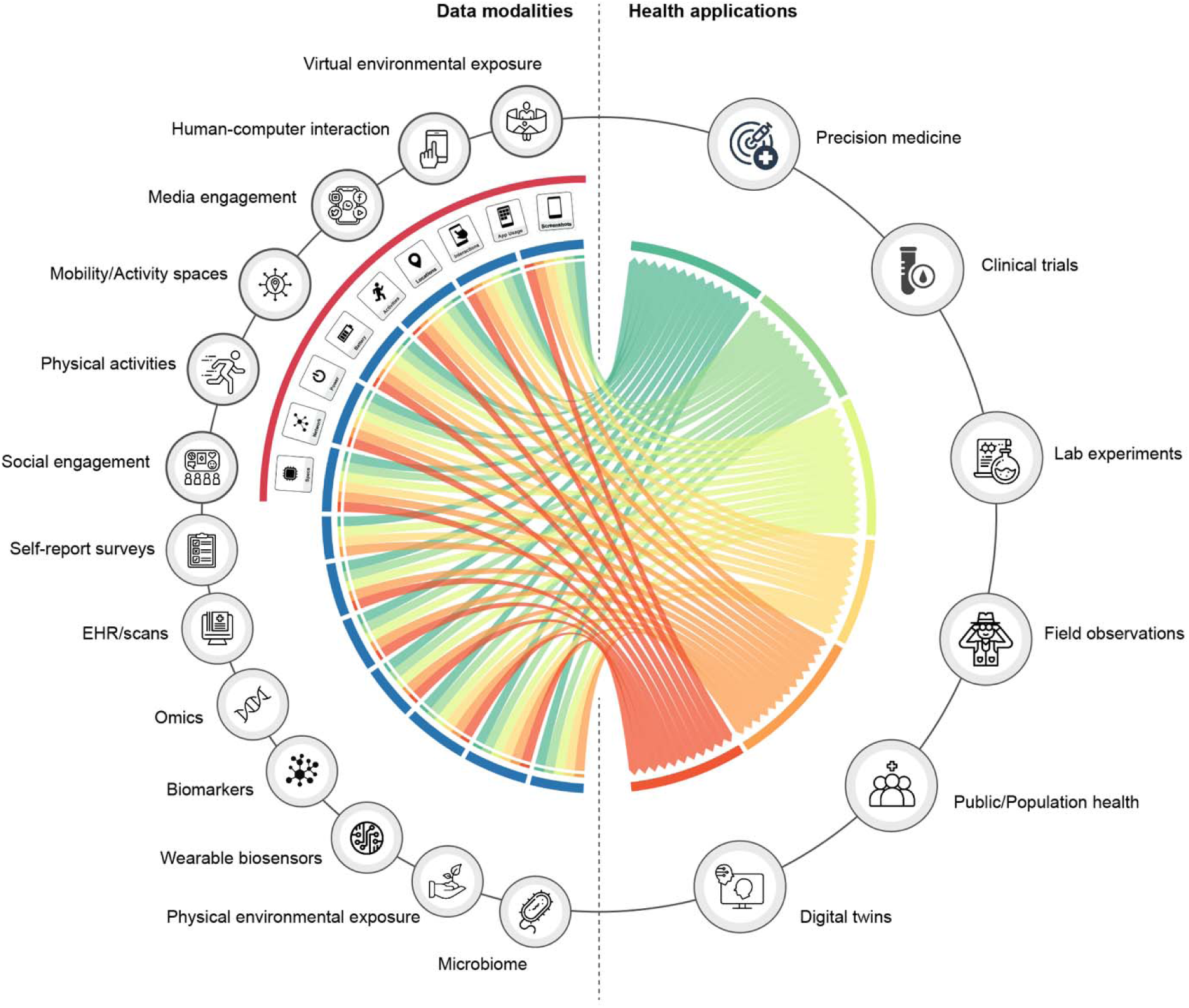
A chord diagram illustrating relationships among multimodal digital trace data, traditional health measures, and research domains. Arrows indicate how different data modalities contribute to various health research and application domains. Square icons denote modules supported by the Stanford Screenomics platform that, individually or in combination, form higher-level digital trace data modalities—such as social engagement and activity spaces—which can serve as key indicators of human behavior and health.

During urgent events such as a pandemic, these data can reveal shifts in physical activity, mobility, and social interactions across populations, providing insights into behavioral adaptations and emerging risk factors, and enabling timely public health responses as situations evolve. Comprehensive digital trace data can also facilitate proactive dose optimization and prediction of potential risks. For example, computational models that integrate multi-layered digital traces with biomedical data can be used to construct digital twins that simulate personalized health trajectories, test interventions before deployment, and guide adaptive, personalized interventions with real-time adjustment. By converting rich behavioral and biomedical data into actionable health insights, comprehensive digital trace data extend research beyond traditional methods, optimize interventions at individual and population levels, and enhance the effectiveness and impact of health applications.

Despite significant interest from research communities, widespread adoption of comprehensive digital traces – as in Screenomics – has been limited due to the lack of tools for the continuous collection of content and interaction data and the computational challenges faced when processing vast collections of unstructured data generated by continuous monitoring. Studies collecting such data have often relied on participant data donations (Ohme et al., 2024; Rodrigues et al., 2018). Sourcing data through donations is often easier to implement and can sometimes capture information unrestricted by smartphones’ operating systems. Much has been learned from these data, but the burdens placed on the participants during data curation may lead to non-ignorable missingness and selective reporting biases. A handful of studies have built tools for passive collection (low participant burden) of Screenomics type data, including possibility to track smartphone users screen interactions (Antal et al., 2015; Zingaro et al., 2024) and capture screenshots of screen content (Yee et al., 2023). Most of these studies relied on custom applications that are not publicly accessible. And, although Yee and colleagues’ (2023) ScreenLife capture was introduced as an open-source solution for researchers’ broad use, the software has not been adopted beyond its developers (and another experienced software developer) and is no longer actively maintained.

As shown in **Table 1**, various commercial and academic offerings have sought to promote smartphone-based digital trace research (Estrin & Sim, 2010; Ferreira et al., 2015; Jardine et al., 2015; Lakshminarasimhappa, 2022; Menaspà, 2015; Onnela et al., 2021; Wang et al., 2017). However, no platform to date supports both quantity-based and quality-based data collection. Many existing platforms are “closed,” and their tendency to focus collection narrowly on just a few sensor streams makes them difficult to scale. Some open-source frameworks (i.e., AWARE, Open Sensing Project, Open mHealth, KoBoToolbox) do exist, but they require substantial technical expertise and resources to customize data collection parameters (e.g., sampling interval, image quality) that align the tools with specific research objectives, making them challenging to use and replicate. In practice, these platforms also function as “closed” platforms because only the original developers can add new features and improvements. Users cannot expand the capabilities of these tools to meet evolving research needs, and in some cases the lack of transparency into the data collection and analysis processes precludes reproducibility. Scientific rigor and validity is compromised when researchers do not have access to the raw data or detailed documentation on how algorithms process and filter information. In sum, the limited customizability, scalability, accessibility, and transparency of currently available tools and frameworks for comprehensive digital trace data collection often poses a significant barrier for researchers.

**Table 1.**
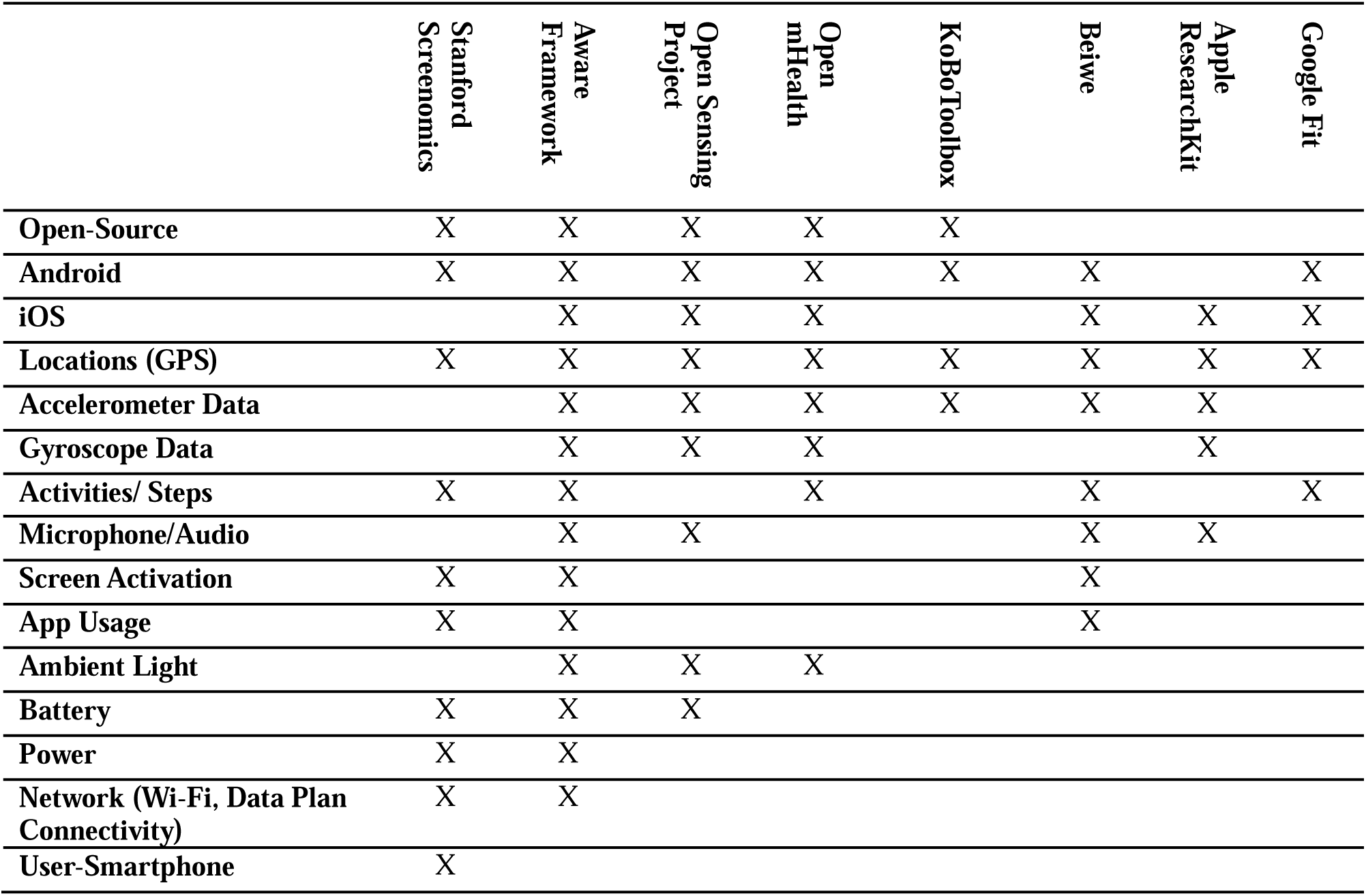
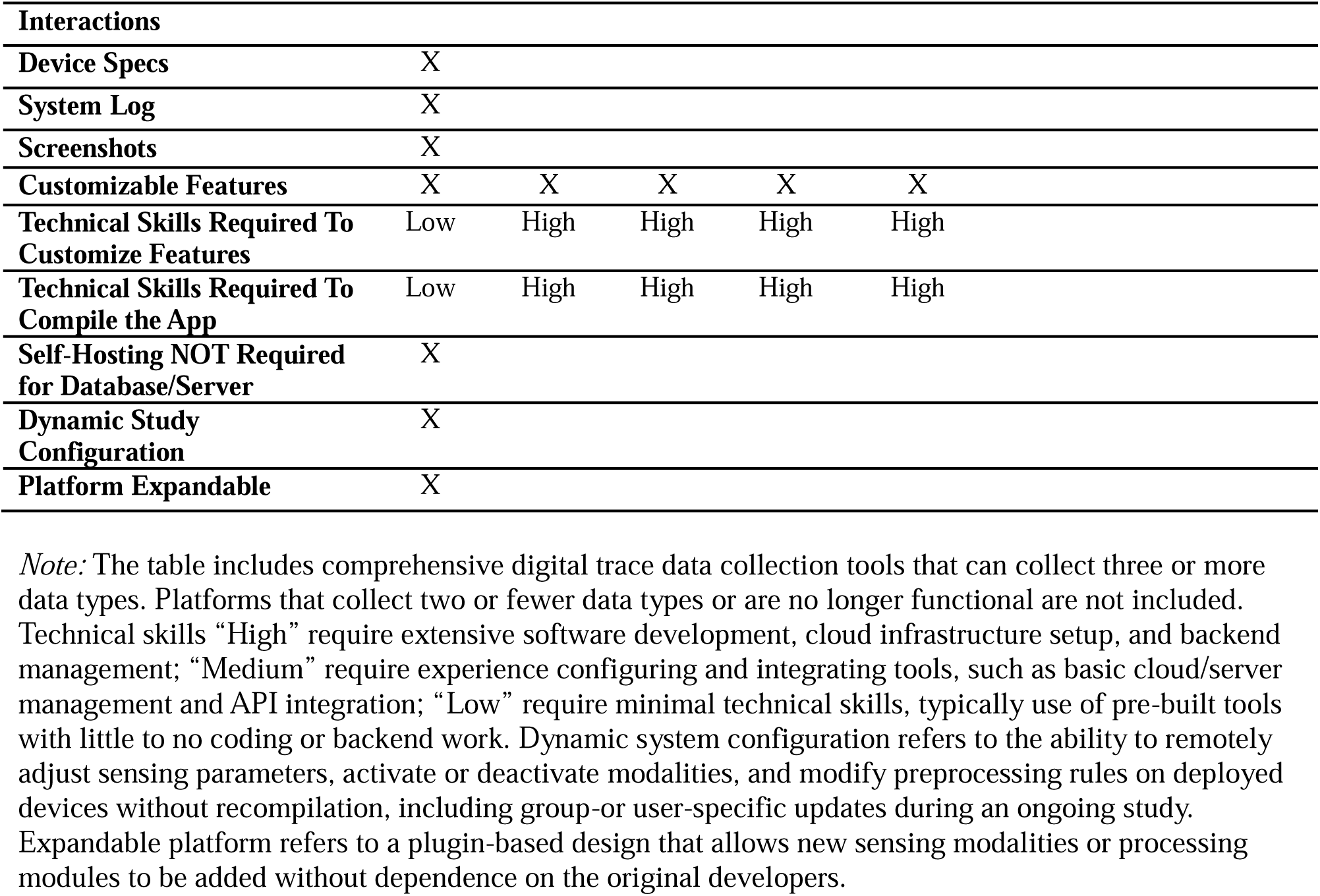
Comparison of Digital Trace Data Collection Platforms: Key Features and Capabilities.

To address these gaps, we developed an open-source smartphone-based digital trace collection platform that is easy-to-use, open-source, and expandable. The Stanford Screenomics platform consists of two native Android applications (apps) and a back-end data repository. The platform’s Data Collection app is easy to deploy, offers extensive customization options enabling researchers to tailor, with minimal additional development, the types of data recorded, sampling frequency, data transfer methods, and upload cadence. Complementary to the Data Collection app, the Dashboard app supports study coordinators’ real-time monitoring of participant data, identification of data collection issues, and automated reactive messaging with participants. The platform’s back-end uses a scalable NoSQL database (Firebase Firestore and Google Cloud Storage) for secure, HIPAA-compliant data storage. The platform is specifically designed to flexibly match specific requirements of many types of studies seeking to leverage the many advantages of continuous, automatic, passive data collection, including increased data completeness, reduced participant burden, minimal reporting biases, and the ability to perform real-time monitoring. In open-source form, the platform supports the reproducibility and replicability of research studies by providing thorough documentation of data collection and processing methods.

As seen in **Table 1**, the Stanford Screenomics platform, with collection of screenshots and user-smartphone interactions, is currently the only tool that allows researchers to match granular records of the actual content that appears on smartphone screens with the behaviors that precede or follow those screen experiences. Particularly useful for study of health is the possibility to align the timestamps associated with each screenshot with behavioral events that occur prior to or after specific kinds of content appear on the screen. For example, content-based matching methods where OCR and image analysis of screenshots are used to detect specific keywords (e.g., “glucose spike”, “stress level”) or pictures (e.g., of people engaged in physical activity) allow researchers to link the screen content to specific behavioral action (e.g., viewing a medication reminder screen in the 10 seconds before logging medication adherence, or examining a physical tracking app screen in the minute prior to a substantial increase in step-counts). Similarly, event-triggered matching uses specific screen or physical actions—such as receiving a fitness app notification, opening a symptom diary app, or crossing a geofenced location near a healthcare facility—as anchors for identifying systematic patterns of linkage between content and behavior. By combining temporal and content cues, researchers can achieve more accurate and meaningful associations between screen data and user behaviors.

Our goal is to support the development of versatile, user-friendly, and highly customizable tools for collecting comprehensive digital trace data, and enable collaborative extension of those tools by the research community. To our knowledge, no existing system offers this comprehensive set of capabilities in a cohesive manner. In developing the open-source Stanford Screenomics platform, our hope is to provide a significantly advanced and easy-to-use tool that both supports and expands the scope and accessibility of digital trace research. The platform is currently built for Android OS. While in principle the platform architecture might also be implemented on iOS, prohibitions in Apple’s infrastructure do not allow for continuous screenshot collection. It is thus not possible to collect all the same data streams on iOS. Acknowledging that, given demographic and usage differences between platforms such as Android and iOS, restricting data collection to the Android operating system may introduce some sample-level selectivity, there is no evidence thus far that the underlying psychological, physical, and behavioral processes manifest differently on the different operating systems. In the following sections, we describe the Stanford Screenomics platform’s architecture, data flow, feature customization, collaborative module extension methods, and data collection and communication. We then demonstrate the utility of the Stanford Screenomics platform for studies leveraging digital trace data with *illustrative 24-hour digital trace data* and point to the range of ways this easy-to-use open-source platform can, together with the community of developers, expand the range and reach of digital trace research. The open-source Stanford Screenomics platform project files, detailed component documentation, and step-by-step instructions with visual guides to support researchers use and developers extension of the platform are available here: https://github.com/iansulin/stanford_screenomics

## METHODS

### Preparing for Screenomics Research

The Stanford Screenomics platform provides a framework that enables researchers to build an Android-based smartphone application (app) to capture and organize smartphone screen content, smartphone-user interactions, and other behavioral data, such as step counts and geographic location, in participants’ natural environments. While no programming expertise is needed, successful studies require careful planning across several key areas: backend infrastructure and compliance, frontend device selection, study design, app compilation and configuration, app distribution, and the overall study timeline. The following sections provide initial guidance on each of these elements to help researchers plan and execute a Screenomics study effectively.

### Backend Infrastructure and Compliance

Given the very personal and sensitive nature of smartphone data, it is paramount that researchers confirm their institutions have infrastructure in place to support ethical engagement with, collection of, and storage of comprehensive digital data trace data. Before beginning such a study, the research team should confirm that the institution’s backend—the server-side systems and services that manage authentication and databases—support and storage capabilities ensure compliance with IRB and security standards. The Stanford Screenomics framework currently relies on Firebase (i.e., Authentication, Firestore, and Cloud Storage) which is HIPAA-compliant and automatically configured during app compilation. Some institutions may support alternative backends, such as Amazon Web Services, Azure, or local servers. The Stanford Screenomics framework does not currently provide automatic setup for these, so a developer may be required to implement custom backend integration. Researchers must confirm with their IRB that the chosen data storage solution meets institutional security, compliance, and ethics standards.

### Front-end Device Selection

A second preliminary consideration is the “front-end” device participants use to supply their data. The data collection app built on the Stanford Screenomics framework is compatible with any Android device, with Android versions 11 or higher recommended for optimal performance. Google Pixel and Samsung Galaxy devices are generally the most reliable. Researchers may allow participants to use their own devices or provide study-issued phones. Letting participants use their own Android phones reduces costs and provides for better capture of naturalistic behavior, but introduces variability in hardware, operating systems, and device configurations that may affect data quality. In contrast, providing participants with study-issued phones ensures consistency in data collection processes and provides for easier troubleshooting, but introduces additional costs and logistics like transferring devices, maintaining data plans, and promoting participant adherence. The data collection app is not available on iOS because of iOS restrictions.

### Number of Participants, Study Duration, Data Volume and Cost Estimation

Knowing that the general infrastructure for comprehensive digital trace collection is in place, researchers can then turn to the details of their specific study. Although apps built on the Stanford Screenomics framework can accommodate studies with any number of participants, data streams/volume, or study duration, cost estimates should be reviewed. Researchers can use the Google Cloud Pricing Calculator to estimate costs for their specific usage patterns (https://cloud.google.com/products/calculator).

With default settings described in the GitHub documentation, capturing about ten hours of smartphone data per day from all nine modules at a smartphone resolution of 1080 × 2400 pixels generates roughly 10 GB of data per participant per day, though data volume can vary across devices. Different modules, data types, or sampling rates can be assigned to study groups or individual participants without coding, which will affect storage and operational costs. For example, under a pay-as-you-go plan (∼$0.020–$0.026 per GB-month as of August 2025), 20 participants generating 10 GB of screenshots per day over four weeks (28 days) would produce approximately 5,600 GB (∼5.47 TB). Assuming linear accumulation, the average stored volume is about 2,800 GB (∼2.73 TB-months), with raw storage costs of $56–$73. Downloading the full dataset at $0.12/GB would cost $670, plus ∼$20 for additional operation fees. Meanwhile, one participant generates ∼2 million Firestore event documents per day; for a 20-participant, four-week study, text-based storage costs are ∼$200–$250, with read/write operations adding ∼$2,000, giving a total estimated cost of ∼$3,000. Reducing screenshot quality to 25% and doubling module sampling intervals (e.g., GPS every 20 minutes instead of every 10) can lower total costs to ∼$750 to $800. In our experience, cost considerations play into key decisions around which and how many Stanford Screenomics platform modules to deploy to get the data (and sampling intervals) needed for answering specific research questions.

### App Compilation and Configuration

Once the study design is known, researchers will need to compile and configure a study-specific Stanford Screenomics app that will be given to participants. Open-source project files available through GitHub (GitHub account is not required for download). Building a custom study app using the Stanford Screenomics framework involves compiling the app in Android Studio software on the researcher’s laptop or desktop computer. Android Studio, which must be installed prior to downloading the project files, can run on Windows, Linux, or Mac, so there are virtually no restrictions on the type of laptop or computer used for compiling and configuring the app for a specific study. While a high-end machine is not necessary, the laptop or computer should be capable of handling app compilation and managing large datasets efficiently. For smooth builds and caching, a modern quad-core processor, 16 GB of RAM, and an SSD of at least 512 GB are recommended.

App compilation prepares the study data collection app and dashboard app, integrating all backend connections without requiring programming skills (step-by-step instruction documentation is available on GitHub along with associated videos). During compilation, researchers finalize data collection modules, sampling rates, and screenshot quality, which directly influence storage requirements and operational costs.

### App Distribution

Once the app is compiled, researchers must determine a distribution strategy. One option is direct APK distribution, where researchers send the compiled app to each participant. This approach allows immediate installation but may raise some security concerns because participants must enable “installation from unknown sources”. Alternatively, the app can be released through the Google Play Store, which provides participants with a familiar installation process, allows researchers to push out automatic updates, and facilitates scalability for larger studies. Google Play distribution requires a developer account, a detailed app listing (e.g., title, description, icon, and screenshots) a public privacy policy describing data collection and security measures, and justification for sensitive permissions such as screenshots or location tracking. The target audience for the study app must be specified, particularly if minors are involved, and IRB approval documentation may be requested. A researcher-defined study-specific password ensures only authorized participants use the app even when published through Google Play. Note that although Google’s review process may take several weeks and reviewers will request login credentials during the review process, the Play Store provides a secure and participant-friendly distribution method suitable for larger or regulated studies.

### Overall Study Timeline

The study timeline typically begins with confirming backend support and compliance, and thoughtfully planning the study design, including participant inclusion criteria, app module selections, and data quality considerations with associated cost estimates. Because approvals from the IRB and Play Store can take substantial time, it is recommended to address these early in the planning process. Researchers may consider preparing IRB-ready materials covering informed consent, participant privacy, withdrawal procedures, and any relevant legal or regulatory requirements. In the U.S., university IRB approval usually takes about two to three months. During this period, the app can be customized, configured, and compiled. Once IRB approval is obtained, the app and supporting documentation can be submitted for Google Play Store review. Following Play Store approval, participant recruitment and data collection can begin and continue for the desired study duration.

#### Overall Architecture and Data Flow of the Stanford Screenomics Platform

The platform architecture consists of three main layers: front-end, middleware, and back-end. A comprehensive overview of the system’s operations, platform architecture and data flow are shown in Figure 2. The ***front-end*** includes the Firebase console, the dashboard application, and the Stanford Screenomics data collection app. The Stanford Screenomics data collection app is an application that study participants download (e.g., from the Google Play Store or from researchers) to their phone. On launch, participants register with their study group code and ID. On successful login, the app retrieves the relevant study settings and initiates data collection. The data are first encrypted and stored locally on the device, then securely transferred in encrypted mini-batches to ***back-end*** Firestore data bases and Cloud Storage buckets hosted on the HIPAA-compliant Google Cloud Platform (GCP). Text data—structured or semi-structured information such as GPS coordinates, time stamps, and sensor metadata—are stored in Firestore, while non-text data—unstructured data types like screenshots—are saved in Google Storage folders. A ***middleware*** module handles authentication, secures communications between the front-end and back-end, and oversees data transfer. User identities and access are verified through Firebase Authentication to ensure secure app use. Privacy and security are maintained across three levels: at the app level with local encryption and secure authentication; at the GCP level with encrypted data in transit and at rest, fine-grained access controls, and compliance with HIPAA; and at the institutional level through customized security policies, monitoring, and adherence to IRB and ethical standards. Alongside the participant-facing *data collection app* and researcher-facing *Firebase console* is a *dashboard app* that facilitates study coordinators’ real-time monitoring of data collection and, if needed, communication with specific participants.

**Figure 2.**
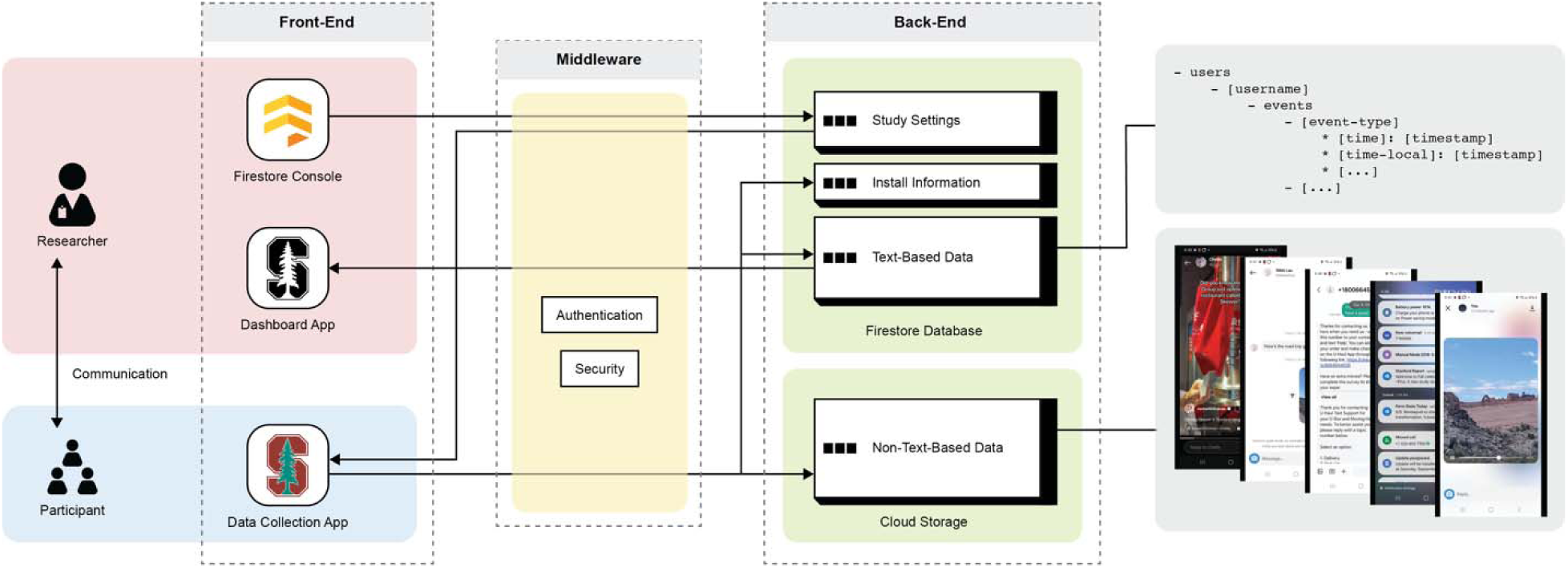
General platform architecture and overall data flow

### Firestore Console

While the Firebase Console is not inherently designed as a front-end system, it effectively functions as one in this platform. Researchers set up and launch a study by interacting with the [`settings_profile`] collection in the Firestore database. Through this console, researchers can assign each participant a study group code (e.g., intervention or control) and configure group-or individual-specific study settings, such as data sampling frequency and sensor activation.

### Data Collection app

The Data Collection app is designed to collect and process the different data types from smartphones. As shown in the Stanford Screenomics column in **Table 1**, current modules facilitate collection of screenshots, app usage logs, user-smartphone interactions, location updates, physical activity data, battery status, network connectivity information, and device logs. Data collection capabilities may be easily expanded through creation of new modules to accommodate additional data types. The app runs continuously in the background, without the need for any human interaction or guidance, as participants use their smartphones normally in daily life. The app starts automatically upon the first login and remains active until logout. If the device is ever turned off completely, the app automatically resumes data collection after a device reboot. Data are collected in hidden local storage, encrypted, and later transferred in batches to the back end, with text data sent to Firestore and non-text data, such as screenshots, uploaded to Google Cloud Storage. The app checks connectivity based on individual-or group-specific data transfer preferences set by the researchers before participant onboarding; for example, setting text data to upload over any network while setting the (often larger) non-text data (e.g., screenshots) to only upload via Wi-Fi.

### Middleware

The middleware structure is essential for security and reliability, focusing on authentication, security measures, and monitoring capabilities. Using Firebase Authentication, the middle-ware module verifies user identities, allowing only authorized participants access to the app and safeguarding sensitive information. Firestore security rules based on user roles define specific permissions. For example, trained researchers may have full access and administrators can view data, while participants cannot access their own data. Firebase rules also enforce data validation to prevent transfer of unauthorized formats into the back-end data base. Integration with Crashlytics provides real-time error reporting and analytics, enabling the research team to monitor app performance and quickly address problems. The middleware interacts with both the front end and back end through well-defined APIs—specific protocols that facilitate secure communication, including RESTful endpoints for data retrieval, user authentication, and data submission. This structured middleware approach enhances data management security and efficiency to protect data while allowing researchers to maintain oversight of participants’ data provision.

### Back-end

The back-end of the platform uses a Firestore NoSQL database with four collections: (1) [`install`] for app installation codes, (2) [`settings-profiles`] for dynamic parameter settings, (3) [users-events] for text-based digital trace data storage, and (4) [`ticker`] for individual-level summaries of participants’ recent activities (data provision). A fifth location, Google Cloud Storage, is where non-text-based data is stored in user-specific folders named by username. Both the Firestore database and Google Cloud Storage folders are HIPAA-compliant.

1. The [`install`] collection stores information about the participants who have installed the data collection app. When users (i.e., study participants) first open the data collection app, the app generates a unique install code that is stored on the local device and uploaded to the [`install`] database. These data indicate which participants have installed the app but have not completed the setup (via cross-reference with study enrollment data), and which participants have dropped out (via cross-reference with `CaptureStartupEvents` in the [`users-events`] database. Currently, these comparisons are done manually.
2. The [`settings_profiles`] collection serves as a centralized hub for configuration settings tailored to various user groups, allowing researchers to dynamically assign or modify parameters throughout the study via the Firebase console. Currently, there are sixteen parameters that can be dynamically controlled to tailor the app to specific research needs, such as user profile refresh intervals and step count sampling frequencies. If a parameter like location-enabled is changed, the app will adjust GPS data collection accordingly.
3. The [users-events] collection organizes data hierarchically under user-specific collections, accessible via unique documents named by following usernames (group code + participant ID). Each event log contains specific structures, such as latitude and longitude for `GPSLocationEvent` and activity types for `InteractionEvent`, along with two timestamps: standard GMT and device time.
4. The [`ticker`] collection logs recent events for each participant, filtering [`users-events`] data to show the most recent timestamps. This log provides researchers with a quick overview of user engagement and interactions, enabling efficient monitoring of participant behavior. The ticker data are sent to the Dashboard app for real-time tracking.
5. The Google Cloud Storage stores non-text data, namely screeenshots, in individual folders with the username. Each screenshot is saved with a file name that includes the group code, participant ID, and timestamps (e.g., `INT_101_20250122134522.jpg`). The file name is logged in the [users-events] collection under the ScreenshotEvent log whenever a screenshot is taken, allowing for easy matching later.

### Dashboard Application

The purpose of the Dashboard Application is to efficiently monitor participant activity and facilitate real-time communication with participants. Upon receiving the [`ticker`] data, the Dashboard application creates priority indicators for inactive participants, flagging those who have been inactive (provided no data) for over 24 hours. This information allows researchers to monitor participants in real-time and identify individuals who may need to be contacted. The Dashboard also supports direct communication between researchers and participants via email, facilitating engagement and support throughout a study.

#### Setting-Up and Launching a Study

### Preparing the App for Deployment

Study setup using the Stanford Screenomics Data Collection App begins with compiling the application according to specific research requirements, such as determining which data to collect through existing module selection and activation. The user-friendly compiling process, in Android Studio, allows researchers to easily configure and compile the app.

The data collection app features a modular architecture that simplifies data collection across nine distinct modules. This modular structure allows researchers to easily add or remove data collection functions through the `ModuleController`, providing flexibility and scalability, as each module can be independently activated or deactivated based on study needs. This aspect of architecture allows the app to be personalized for any given study. In practice, researchers download the open-source project files of the app and open them in Android Studio, which is available for free. Within Android Studio, the Module Controller can be located in the left-side interface list view, and it serves as the central activation/deactivation interface, listing all available data collection modules (currently nine) with Boolean values: `true` (activated) or `false` (deactivated). Before compiling the app, researchers can set the Boolean values according to their specific study requirements for data collection. The activation or deactivation of a module does not impact the overall performance of the app, as each module operates independently. Some features, like step count and GPS data collection, can be dynamically turned on or off throughout the study, allowing a single app version to accommodate changes in data collection. However, once a module is deactivated, it cannot be reactivated later, thereby requiring separate app compilations for each study group.

Additionally, to prevent device storage overload, the app automatically restricts screenshot capture when the user device’s available storage falls below 50 MB. This architecture ensures that researchers can efficiently customize the app for their studies without extensive technical expertise and participants can simultaneously use their smartphone as normal while providing rich multimodal data.

In addition to activating modules, essential configuration steps—such as setting up an app password and defining a consent form—can be easily modified within two dedicated configuration files, eliminating the need to scroll through code to find placeholder text. The consent form serves as a prominent disclosure, required by Google’s User Data Policy for Android apps that collect personal and sensitive information (https://developers.google.com/terms/api-services-user-data-policy). The Google policy mandates that developers clearly inform users about data collection practices and obtain their informed consent. It is important to note that Google’s consent form must be added to, and may not replace, Institutional Review Board (IRB) consent forms. Therefore, two consent forms are generally required: one for Google policy compliance and one for ethical research approval through an IRB.

Researchers must also configure Firestore, Google Cloud Storage, and Crashlytics by registering the app in a new Firestore project. The automatically generated JSON file should then be placed in the main app and the database manager modules of the Stanford Screenomics app using Android Studio. This drag-and-drop action automatically sets up Firestore, Google Cloud Storage, and Crashlytics. Finally, researchers need to ensure that Crashlytics is enabled in the Firebase for their new project. Not doing so would prevent the app from capturing and reporting crashes or errors, making it difficult to maintain reliability and improve user experience. Optionally, researchers can also manually configure Firestore rules for data security.

Once all configurations are finalized, compiling the app is straightforward. Researchers select “`Build`” from the menu in Android Studio, then choose “`Build Bundle(s)/APK(s)`” and “`Build APK(s)`,” after which a notification will confirm that the APK (Android Package Kit) file has been generated as a ready-to-use installation package for Android devices. To distribute the APK to participants, researchers can share it via email, cloud storage links, or secure file-sharing methods. Participants can then install the app by downloading the APK file onto their Android devices, opening the APK file and following the on-screen instructions. Thorough testing should confirm that the app meets the study’s specifications, ensuring participants have access to the latest features and updates.

### Dynamic Controller Parameter Setup

Prior to onboarding participants, researchers can customize the data collection app’s features to align with specific study requirements, including adjustments to sampling intervals and data transfer network preferences. The app periodically retrieves the latest parameter values at intervals specified in the settings profiles (through a dynamic parameter called ‘`settings-refresh-interval`’), allowing researchers to update data collection methods as needed, without needing to recompile the app or contact participants to makes changes. This dynamic control allows researchers to balance the frequency of updates with considerations for device battery life, storage, privacy, and study needs. Currently, sixteen features can be dynamically controlled to tailor the app to specific research needs. There are no fixed default parameter values, as these can vary widely depending on the number of activated modules. For example, a 5-second sampling interval for GPS coordinates may not impact performance if only the location module is active. However, if combined with a screenshot module set to capture every 5 seconds, this frequency could strain system performance or rapidly drain the battery. The `settings` section in our GitHub documentation provides recommended minimum parameter values (highest resolution) for a case where all nine modules are activated. For researchers interested in detailed, module-and device-specific performance estimates, including battery consumption, RAM usage, CPU load, and storage requirements, a `pre-module activation guide` is available under the `App Compilation`. Despite varying demands across modules, with screenshots being most intensive and GPS notably impacting battery, differences observed across devices were minimal.

When the data collection app is configured with the `google-services.json` file, the app automatically creates four collections in the Firestore console: [`install`], [`settings-profiles`], [`users`], and [`ticker`]. To set up dynamic controller parameters, researchers must first create a new document named “`_default_`” under the [`settings-profiles`] collection. In this document, researchers must add all parameters from the full list of available parameters for the app version and set default values for each parameter according to the study needs. The data collection app imports these default values during a participant’s first login. If all parameters are not specified in the “`_default_`” document, the app will lack reference values, potentially hindering its ability to collect data effectively and impacting overall study outcomes. Therefore, it is crucial to finalize these settings before onboarding participants.

Researchers can create as many additional profiles as needed for different study groups by adding group codes, similar to how the default profile is set up. For example, if participants have varying data transfer preferences, two profiles can be created with the group codes [`WIFI`] for those who prefer transferring data over Wi-Fi only and [`DATA`] for those willing to use their mobile data plan. These group profiles do not need to include all parameters from the list; only those that are relevant need to be specified. Researchers can configure data transfer settings differently for each group while keeping other study parameters consistent. This allows for tailored data collection strategies based on the specific requirements of each group. The group codes are later assigned to participants who will use them along with other information provided by researchers to register in the app during onboarding. If a participant registers in the app with a group code that does not match any created profiles, they will inherit the default setting parameter values specified in the “`_default_`” document.

### Participant Onboarding

To initiate a study with the Stanford Screenomics Data Collection App, researchers must first establish participant groups, such as intervention and control groups. Each participant requires a unique group code (e.g., “`WIFI`” for the over Wi-Fi only group and “`DATA`” for the unrestricted data group), an individual ID number (e.g., `101`, `102`), an email address, and a password for registration and login. The password should be configured during the app’s setup before compilation. While researchers provide the group codes, individual ID numbers, and password, participants must also supply their own active email addresses during onboarding. The email is required for Firebase authentication, restricted database access, and researcher-participant communication via the Dashboard app. However, email addresses are stored only within the Firestore authentication system and are not linked to study data, and they are not visible within the app, database, or storage. All collected data are associated with a unique username, composed of a group code and individual ID numbers, ensuring that study data remains non-identifiable through email.

Upon first login, participants will see a one-time pop-up prompting them to review and sign the Google permissions consent forms. If they agree, they will then be asked to grant necessary permissions for the app to function effectively, such as access to precise location data and media projection features, depending on the modules activated during app compilation. Once these permissions are granted, data collection begins passively in the background, ensuring an unobtrusive experience. Screen recording is continuous unless participants log out of the application or choose to pause screen recording. The app’s main interface will display a single button for participants to pause or resume screen recording at any time, allowing them to manage storage, battery, and privacy concerns. To ensure continuous data collection, the app sends reminder notifications if a participant pauses the screenshot collection: one message after 30 minutes, and then at 7 AM and 7 PM for the next two days, totaling five notifications. If participants do not agree to the consent or fail to provide all required permissions, the app will not advance to data collection. Consent and permission requests will not reappear on subsequent logins unless permissions have been revoked. If participants revoke any permissions during data collection, the app will pause that specific data collection. Upon their next app launch – whether by actively opening the app, rebooting the device, or tapping a notification sent after 30 minutes of pausing – they will be prompted to re-grant the revoked permissions, ensuring the study remains compliant with ethical standards.

### Monitoring of Data Flow

Effective monitoring of data flow is essential for maintaining the integrity and reliability of research outcomes. By continuously tracking participant engagement and data collection processes, researchers can make informed adjustments, address potential issues in real time, and enhance data quality. In the Stanford Screenomics platform, two primary methods are employed for monitoring data flow: the [`users-events`] collection and the [`ticker`] collection in Firestore. The [`users-events`] collection provides detailed insights into individual participant actions, while the [ticker] collection offers a streamlined overview of user engagement.

The [`users-events`] collection captures a range of text-based event data. An event denotes a specific action that occurs at a particular moment, encompassing various user activities such as app logins and logouts, pausing or resuming screenshot functions, and interactions with notifications (e.g., read or ignored). It also includes all text-based event data captured by activated modules, as well as device system and app lifecycle events, such as device reboots and automatic app launches. Events are organized by type and timestamps, often accompanied by additional metadata specific to each event. For non-text-based events, a dual approach is employed: when an event occurs, the actual data—such as screenshots— is sent to Cloud Storage, while simultaneously, the [`users-events`] collection logs that the event has taken place, including corresponding timestamps and metadata such as the file location URL in Cloud Storage. This approach allows researchers to track both non-text-based and text-based data within the same structure and in one location. Consequently, this alignment facilitates not only the quick identification of trends but also enables comprehensive in-depth analysis later.

The [`ticker`] collection filters the [`users-events`] collection data to create individual user documents, which list each event along with the most recent timestamp. This collection is particularly beneficial as the number of app users increases, as manually checking each participant’s events for recent activity can become labor-intensive. When a participant’s app syncs their events to the [`users-events`] collection, the corresponding fields in the [ticker] collection are updated to reflect recent activity across different event types. This [`ticker`] data is organized and displayed in the Dashboard app, allowing researchers to easily view all study participants’ usernames ranked by the length of inactivity on the main interface of the app. Users who have been inactive across all events for more than 24 hours are highlighted in red. Each username is expandable on tap, revealing a detailed view of all events and the most recent activity timestamps. Researchers can also initiate email communication with participants directly from this view if needed.

#### Platform Extension

The Stanford Screenomics Data Collection App is designed with a modular architecture that facilitates platform extension through easy plug-in integration of new functions, while allowing developers to activate or deactivate existing modules. Currently, the app includes two core base modules—`ModuleManager` and `DatabaseManager`—and nine distinct data collection modules: screenshots, app usage logs, user-smartphone interactions, location updates, physical activity, battery status, system power, network connectivity information, and device specs logs. The core base modules provide essential functionalities that form a robust foundation for each data collection module’s operations. This architecture simplifies the integration of new modules by enabling them to easily adopt the tools and frameworks from the base modules, allowing developers to ensure their custom data collection modules align seamlessly with the overall app requirements regarding data structure, storage setup, network preferences, and data transfer methods.

### Base Module 1: Module Manager

The Module Manager serves as the central control system, overseeing the operational status of various data collection modules. Key functionalities include centralized event record management and timestamp definition, which together create a cohesive environment that enhances consistency in logging and data management. All modules in the Stanford Screenomics Data Collection App follow a uniform data structure within Firestore, storing data under a user’s collection as a list of event documents. Each event in this list is organized by event type and includes two timestamps—one for standard time zone and one for the user device’s system time (usually in the user’s local time zone)—along with a unique ID and additional event-specific metadata. The event record management and timestamp definition functionalities facilitate this organization. Additionally, the `ModuleManager` provides a collection of reusable components (e.g., classes, methods) for various features. For example, a method for reading battery state changes can be employed in the location module to pause data collection when the battery level drops below a critical threshold, conserving energy. In the battery status module, the same method can log battery status at predefined intervals. This capability to share reusable components across modules streamlines development, promotes efficiency, and provides insights without redundant code, enabling new data collection modules to quickly adopt and integrate these functionalities. Lastly, the module controller within the `ModuleManager` allows developers to turn on or off existing modules without impacting overall app functionalities.

### Base Module 2: Database Manager

The `DatabaseManager` is essential for data storage and retrieval, serving as the primary repository for data transfer preferences. It includes comprehensive functionalities for local and external storage setup, network configuration, data syncing, and security, all defined as reusable components. The app stores data collected from each module locally on the device until an internet connection is available. Once online, text-based data is transferred to Firestore and non-text-based data to Cloud Storage in batches, with transferred data automatically deleted from local storage to free up space for ongoing data collection. The local and external storage setup functionality manages this process seamlessly when used in a module. Regarding data transfer, each module allows researchers to set preferences for restricting transfers to Wi-Fi only or allowing any internet connection, using dynamic parameters. The network configuration functionality retrieves these parameters from the Firebase console to make data transfer decisions based on network preferences. To ensure data in Firestore and Cloud Storage remains current, a regular data syncing process periodically checks local storage. Additionally, the security functionality oversees storage solutions for sensitive information, such as authentication details, throughout each process.

### Module Dependencies

To support the platform’s extension through the addition of new modules, it is essential to understand its dependencies which can be categorized into three types: modular, internal, and external. Modular dependencies consist of distinct functionalities imported from base modules, organized into separate data collection modules. For example, text-based data collection modules utilize the `TextBasedEventData` class from the `ModuleManager` base module, while non-text-based modules depend on `NonTextBasedEventData` class, both requiring `EventTimestamp` for consistent timestamp formats. Internal dependencies include classes and libraries from the Android framework and Java standard libraries, essential for basic app functionality such as component management and communication operations. External dependencies, often optional, include third-party libraries that enhance capabilities beyond the Android SDK, such as advanced UI components or improved networking features.

The development process for specific modules illustrates these dependency types in action defining its purpose and establishing a clear structure. For instance, when developing a “Location Data Collection” module that periodically collects GPS coordinates, internal dependencies such as `android.Manifest` for location access permissions, `android.os` for system interactions, and `java.util` for data structures are crucial for foundational functionality. The module also relies on the Google Play Services location API as an external dependency for accurate GPS tracking, enabling the collection of real-time longitude and latitude data. Lastly, modular dependencies like `ModuleController` class from `ModuleManager` base module enable the activation or deactivation of individual data collection modules, while `EventTimestamp` class ensures consistent timestamping of each GPS coordinate log. The `ModuleCharacteristics` class standardizes the structure of location event data, capturing key details like class name and event type. The `EventOperationManager` class from `DatabaseManager` base module processes this standardized data. It formats the data, buffers it in memory, stores in local storage, and performs batch uploads to Firestore using `EventData.Builder`. Additionally, `DatabaseManager` base module provides space researchers to add dynamically parameters through `SettingsManager` class.

In contrast, when developing a Screenshot collection module, the process shares similarities but includes unique requirements. This module would integrate additional internal dependencies such as `android.graphics` for bitmap manipulation and `android.media` for screen capture through the `MediaProjection` API. While this module does not require external dependencies, it needs additional modular dependencies like `NonTextBasedEventData` class from the `DatabaseManager` base module to manage and transfer of screenshot images to Google Cloud Storage. Figure 3 illustrates module dependencies of the Stanford Screenomics Data Collection platform.

**Figure 3.**
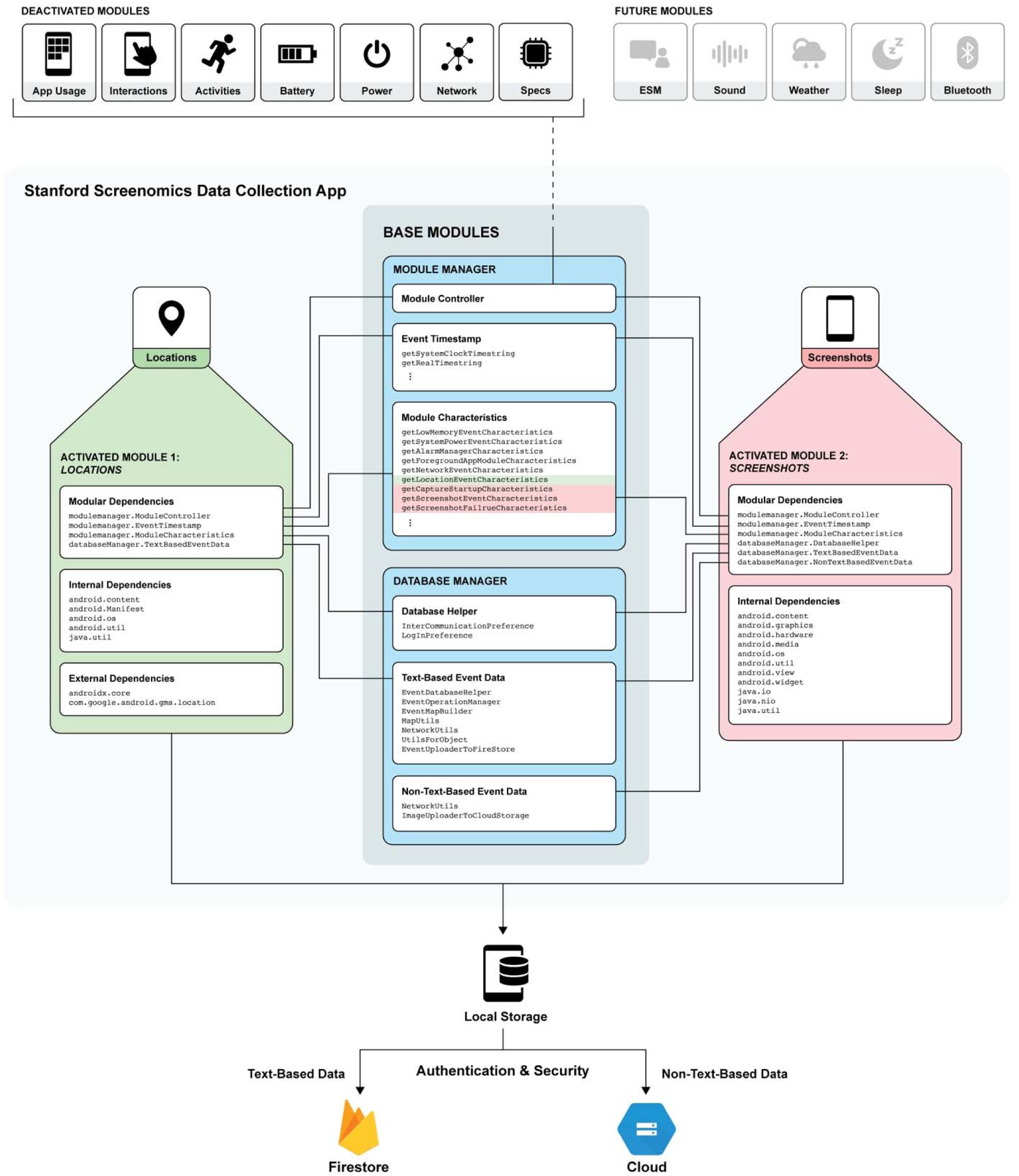
Illustrative Diagram of Stanford Screenomics Data Collection Module Dependencies

## RESULTS

### Illustrative Data

We demonstrate the utility of Stanford Screenomics platform for digital trace research with *illustrative 24-hour digital trace data* collected by the developer (IK) using a Samsung Galaxy S21 smartphone. These data were only collected for demonstration of the platform and app functionalities. The data are not meant to provide generalizations about human behavior and thus do not constitute human subjects research.

#### Activated modules and parameters

The Data Collection App was compiled with all nine currently available collection modules: (1) the *Screenshots* module captures screenshots at defined intervals, saves the screenshot images, and logs information about the screenshot taken (e.g., image file name and screenshot timestamp), (2) the *Apps* module logs the currently active application by checking the foreground app periodically and recording changes with timestamps, (3) the *Interactions* module tracks user events such as scrolls, clicks, and gestures through accessibility services, (4) the *Locations* module logs latitude and longitude from device’s GPS sensor at set intervals, pausing when no updates are available, (5) the *Activities* module tracks user step counts at fixed intervals, (6) the *Battery* module monitors battery levels (low below 15%, okay above 20%) and charging events, (7) the *Power* module logs changes in screen state, noting whether activation is due to user interaction, (8) the *Network* module tracks connectivity changes (Wi-Fi and cellular), and (9) the *Specs* module collects basic device specifications upon account creation. The Screenshots, Locations, and Activities modules log data at researcher-defined intervals, while the other modules record data based on specific events. Data collection interval parameters were set to 5 seconds for screenshots, 1 minute for locations, and 5 minutes for aggregated step counts.

#### Technical performance of platform

Generally, developers try to keep the size of complex app APKs under 50 MB. Here, the compiled app’s APK size of 10.56 MB is well within acceptable limits given that this is a modular app with a total of 11 different modules. Cache size of 67 KB is excellent and indicated efficient storage management. Even with continuous data logging, the app’s impact on device performance was negligible. Generally, keeping RAM usage under 200 MB during active use is essential for smooth performance. During the 24-hour data collection period, the Stanford Screenomics data collection app’s RAM usage peaked at 105 MB (approximately 1.28% of the total available RAM). Generally, CPU usage should remain below 15% to ensure responsiveness. Here, CPU usage averaged 2.1% during active sessions. Typically, less than 5% battery drain per hour is considered optimal, allowing for a target screen on time of at least 6-8 hours. The app drained approximately 1% per hour during active use, indicating minimal power consumption. It is unlikely to significantly impact battery life or be noticeable to users during regular smartphone use. Overall, our testing results indicated that even when collecting high-density data (e.g., location interval of 1 minute) the app performed well, storage management was efficient, and the app’s consumption of resources remained within reasonable limits.

Although researchers do not have direct access to these metrics, there are several ways to check for potential issues, in addition to the Crashlytics and Dashboard activities mentioned previously. Researchers can refer to the `ScreenshotUploadEvents` document in the user-events collection of the Firestore database. First, the remaining-bytes field indicates the available storage space on the user’s device, measured in bytes. While not directly related to uploading, this information helps monitor whether users are running out of space. Second, the error field provides a `yes`/`no` value indicating whether the specific upload event encountered an error. Any `ScreenshotUploadEvent` suggesting an upload did not complete will show “`yes`” as the error value.

#### User experience

Across the 24-hour period, there were no noticeable impacts on device use. As intended, the app operated unobtrusively in the background, and did not interfere with the user’s daily activities. There was no noticeable difference in battery drain, no storage issues, and no app performance problems. The Android system performance remained consistent, with no delays, lag, or interruptions.

In line with the intended design, the user was fully informed, via an informed consent process that users engage when they first open the app and register with their researcher-assigned study group credentials and ID code, about what data were being collected and that they had the ability to stop and start the collection at any time. Along with the institutional IRB-protocols required for Screenomics studies, Google’s clear and transparent consent procedures further educate users about the data collection procedures and empower them to make informed decisions about their participation on an ongoing basis. While formal audit reports or usability testing have not yet been assembled, informal debriefs from our prior and ongoing Screenomics studies (> 500 participants) indicate that users are aware that their smartphone data are often being used by corporations (and sold to third-parties), actively appreciate, understand, and engage with study-specific consent processes, and are often specifically eager to contribute their data to university-based research that is separate from corporate interests. Across studies, we find that 8 to 15 percent of participants choose to withdraw during or after onboarding once they learn and consider the details of the data collection (an indication of informed decision-making). Unlike many of the commercial apps collecting digital trace data, the Stanford Screenomics platform purposively allows users to retain control of data collection and data, including the ability to pause/resume screenshot captures at any time, select precise or approximate (1-2 mile boundary) location sharing, and to temporarily halt all data collection by logging out. Although we have not yet found empirical evidence of systematic participant reactivity during data analysis, some participants do report heightened awareness of monitoring that led them to suspend data collection in specific contexts (e.g., during financial transaction, when texting close others; see also Yee et al., 2023). In sum, beyond this specific illustrative deployment, our on-going studies underscore the value of the privacy controls and protections that are built into the core of the Stanford Screenomics platform.

#### Data Summary

A summary of the types and quantity of data obtained through each of the nine modules during the 24-hour observation period are shown in **Table 2**. The total size of the text-based data was 6.08 MB. The total size of the non-text-based data (i.e., screenshots) was 604.15 MB. The Screenshots module collected 1,729 screenshots that chronicled (at 5 second intervals) visual content that appeared on the smartphone throughout the day during approximately 144.5 minutes (∼ 2.5 hours) of screen time. Simultaneously, the Location module logged 1,440 GPS coordinates and the Activities module recorded a total of 1,626 step counts (in 193 5-minute intervals). The Apps module captured 62 changes among 25 unique foreground applications (counted based on unique app package names logged as metadata for each event). The Interactions module logged a total of 2,455 user-device interactions, the majority of which were “`clicks`” (1,363 simple taps on the screen) that indicate button presses or typing on a keyboard (e.g., when texting or emailing). The Battery module recorded one low battery alert at 10% and one charging event, while the Network module documented a total of 48 changes in internet connectivity. During the 24-hour period, the data were periodically transferred in small batches from local storage to Firestore and Cloud Storage when the device was connected to Wi-Fi (60 successful upload events in total recorded by the app package itself, not through a specific module). Overall, the smooth, unobtrusive, and uninterrupted data collection produced a rich multimodal data that is then easily processed and examined.

**Table 2.** Summary of Digital Traces Collected during 24-hour Study Period.

| Module | Data Collected | Quantity | Interval/Frequency |
| --- | --- | --- | --- |
| Screenshots | Screenshots captured | 1,729 | Every 5 seconds |
|  | Screenshot data collection paused | 0 | Upon user-initiation |
|  | Screenshot data collection resumed | 2 | Upon user-initiation or after reboot |
| App Usage | Foreground app changes | 45 | As app transitions occur |
| Interactions | Scroll-up | 785 | As interactions occur |
|  | Scroll-down | 162 | As interactions occur |
|  | Scroll-left | 69 | As interactions occur |
|  | Scroll-right | 64 | As interactions occur |
|  | Clicked | 1,363 | As interactions occur |
|  | Long-clicked | 12 | As interactions occur |
| Locations | GPS coordinates (latitude/longitude) | 810 | Every 1 minute |
| Activities | Step counts | 193 | Every 5 minutes |
| Battery | Battery state: low (<15%) | 1 | System alert |
|  | Battery state: okay (>20%) | 1 | System alert |
|  | Device connected to a charger | 1 | As charging status changes |
|  | Device disconnected from a charger | 1 | As charging status changes |
| Power | Off | 1 | As system state changes |
| Network | Connected to Wi-Fi | 24 | As connectivity changes |
|  | Disconnected from Wi-Fi | 3 | As connectivity changes |
|  | Connected to data plan | 20 | As connectivity changes |
|  | Disconnected from data plan | 1 | As connectivity changes |
| Specs | Device specifications | 1 | Upon account creation |

## DISCUSSION

Digital trace data collection platforms offer a possibility to further develop and expand the range of insights obtained from diverse research projects across multiple disciplines. A growing body of evidence highlights the value of digital trace data in monitoring and detecting various social, behavioral, cognitive, affective, physiological, and environmental changes, providing deeper insights into social ecology and ultimately improving the management of various associated health conditions (Freelon, 2014; Huckvale et al., 2019; Insel, 2017; Jungherr et al., 2017; Liang et al., 2019; Ohme et al., 2024; Thylstrup, 2019). The proliferation of personal smartphone devices, coupled with advancements in technology, has enabled unprecedented fine-grained and objective data collection (Cheng et al., 2017). However, as shown in **Table 1**, very few of the currently available data collection tools allow researchers to curate their studies effectively due to challenges including restricted scalability, limited customizability, lack of transparency, and the substantial technical expertise required to tailor the tool to the specific requirements of a study, such as software development, data engineering, and knowledge of cloud infrastructure—that are rarely possessed by many behavioral, social, and medical researchers.

### Stanford Screenomics platform

To address these gaps, we developed and introduce here an easy-to-use open-source platform that enables in-situ, real-time capture of multimodal digital traces from users’ smartphones as they go about their everyday lives. Building on the initial Stanford Screenomics app introduced in 2019 (Reeves et al., 2021), the new Stanford Screenomics platform provides a robust framework that allows researchers to capture and analyze the full array of digital life experiences that flow across and through individuals’ smartphones. Extending and integrating existing solutions into larger-scale clinical or research systems usually requires extensive modifications to the underlying architecture of data collection tools, which can hinder research adaptability and responsiveness to evolving study needs. Earlier research has indicated the importance of a flexible framework that accommodates diverse research methodologies and data collection strategies. This expanded framework enables researchers to explore user behaviors across multiple time scales, investigating how moment-to-moment exposure to digital content influences long-term patterns such as content-switching behaviors, technology addiction, physical activity, emotional well-being, and how personal characteristics shape digital engagement, thereby greatly enriching our understanding of the interplay between technology and human behavior and health. The platform has now evolved into a comprehensive digital trace data collection tool by integrating a broader array of data types beyond just screenshots and app usage, offering several significant advantages.

### Technical Advantages

The modular architecture design used in the new Stanford Screenomics platform allows researchers to tailor their data collection processes without being constrained by rigid application structure. The modular approach not only enhances the system’s extensibility but also enables anyone to use partial functionality required for their specific use case. The Stanford Screenomics platform further applies this modularization to encourage community contributions by providing reusable solutions that enable developers to create new modules and expand the platform’s capabilities through well-defined extension points, and allowing for free sharing of select functionalities. This approach also facilitates co-maintenance of transparent high security and privacy standards. Our goal is to create a robust ecosystem where researchers can efficiently customize their tools for specific studies while ensuring compliance with established data collection and privacy standards. We plan to introduce more data collection modules over time to enable the collection of different digital trace data sources, including Bluetooth connections and audio samples. We also hope to have an iOS version of our platform alongside the Android version, although iOS currently restricts some features, such as screenshot data collection, making them impossible to implement.

### Technical Challenges and Open-Source Extension

While the modular approach provides flexibility in feature customization and an easy solution for utilizing selected functionalities in the Stanford Screenomics platform, it also presents several challenges related to device dependency. The app’s performance is significantly influenced by the specific capabilities of the devices on which it operates. Frequent updates to the Android operating system can lead to changes that impact app performance, such as new permissions or modifications in how features are utilized, potentially disrupting data collection processes. To address this, our core development team actively monitors Android release notes, developer previews, and security advisories, conducting timely compatibility testing on both emulators and physical devices. This process enables early detection of issues and the release of prompt patches or updates to ensure continuity of functionality. Despite extensive testing, performance can vary greatly due to individual device specifications, such as processor speed and sensor accuracy, with older devices often struggling to manage simultaneous data collection from multiple modules. This variability may result in inconsistent user experiences and affect user engagement. Furthermore, multi-module apps tend to be more resource-heavy than single-module apps, potentially leading to increased battery consumption and reduced performance. As new devices are released, compatibility issues may also arise, necessitating ongoing updates to maintain functionality across a diverse range of devices. To ensure sustainability beyond the core team’s efforts, we have adopted an open-source model with a structured approach to collaboration. We maintain a public GitHub repository with clear contribution guidelines, coding standards, and review protocols to help researchers and developers propose new features, submit fixes, and report issues. A public discussion forum and dedicated issue tracker foster open dialogue, while regular release notes and updates keep contributors informed. Although we anticipate the community will play an important role in extending and maintaining the platform, the core team will remain actively involved in reviewing contributions, managing releases, and safeguarding compatibility. We are also exploring partnerships and potential funding sources to provide long-term institutional or grant-based support. Inspired by the success of projects like WordPress (www.WordPress.com), which evolved into a leading open-source platform through extensive community involvement, the Stanford Screenomics platform seeks to foster a long-lasting collaborative ecosystem that supports ongoing maintenance, innovation, and research integrity.

### Research advantages

The new Stanford Screenomics platform expands a variety of research possibilities. First, by combining multimodal data sources, researchers can cross-validate findings, improving the reliability and robustness of conclusions compared to relying on a single data type. For example, in a study exploring the relationship between social media content consumption and mental well-being, incorporating app usage data and GPS location data alongside screenshots (which capture content displayed) would enable researchers to assess whether changes in mental health correlate with specific locations, certain app functions and/or particular content. This cross-validation would not only enhance the reliability of conclusions but also would help rule out confounding variables, ensuring that observed effects on the outcome of interest are genuine.

Second, the integration of multiple data sources would allow for the application of advanced analytics—such as machine learning or predictive dynamic modeling—which could uncover complex interactions between often intertwined domains across multiple time scales and contexts. These advanced techniques offer more nuanced interpretations of how contextual and environmental factors influence both momentary and long-term behavioral and health outcomes. Such analyses would be impossible to apply with only one or two data sources; the richness of multimodal data opens new opportunities for more comprehensive insights.

Third, with access to screenshots and user-smartphone interaction data adding to other smartphone use and behavioral metrics, researchers can now identify complex multidimensional, digital phenotypes that offer deeper insights into individual’s life stages and transitions–from second-by-second changes in screen experiences to characterizations of screen experiences over days, weeks, and even years. For instance, while screenshots may reveal that a person has shared on social media their goal of achieving 10,000 steps daily as part of a fitness regimen and subscribes fitness models’ feed, step count data can confirm whether they are meeting that goal, and GPS data can indicate whether they are engaging in indoor or outdoor activities. This multi-dimensional approach provides a clearer picture of the person’s motivation and behavior, creating a richer, more comprehensive individual profile. Finally, the granular real-time digital trace data combined with advanced analytics, enables the design and delivery of personalized, adaptive interventions. For example, if a person’s social media posts suggest an interest in achieving daily step goals but their step count data repeatedly shows low engagement on Mondays while playing games, targeted content—such as motivational messages or goal reminders—can be delivered to encourage action every Monday when starting game play. These insights allow dynamic adjustments to intervention strategies based on changing patterns or participant feedback, ultimately improving behavioral and health outcomes. This is particularly valuable in longitudinal intervention studies, where researchers can adapt interventions to optimize impact and sustain positive behavior change.

### Ethics

While the platform offers valuable insights for advancing research and improving health outcomes, its ability to capture highly detailed personal information and digital trace data creates a risk of misuse by bad actors. The data collected through the Stanford Screenomics platform could be exploited for purposes such as surveillance, targeted manipulation, or unauthorized commercial use, potentially leading to violations of privacy, coercive cognitive or behavioral modifications, or even social control. To mitigate these risks, the research community must take proactive measures to ensure ethical use. This includes implanting robust data privacy protections, obtaining informed consent, and establishing clear boundaries around data sharing (Maeckelberghe et al., 2023; Nebeker et al., 2020, 2021).

As an open-source project, we recognize the challenges in controlling access to and the intended uses of publicly released code. The approach taken here is that active support of independent researchers that are separated from commercial sectors and seeking to collect and analyze rich digital trace data is essential for advancing science and protecting the public interest. In the absence of open-source tools, data collection is dominated by companies with limited transparency and oversight. From this perspective, open access provides an ethical, regulated alternative that reduces misuse risks. The general idea is that benefits obtained through support of external audit, ongoing improvement, and scientific progress will outweigh the (perhaps already happening) misuse risks.

To mitigate the risk of unauthorized collection of sensitive data, we implement an education strategy to support informed consent and user awareness for both platform users (“researchers”) and app users (“participants”). Stanford Screenomics platform licensing specifically emphasizes the sensitivity of screenome data and outlines privacy and confidentiality protocols. The use license highlights that the collection of participants’ screenomes represents a rich repository of digital life, and requires careful attention to privacy and confidentiality. While the app code is publicly accessible, data collection requires Firebase email/password authentication, explicit Android system permissions that mandate credential exchange between researchers and users, and Google Play Store distribution with IRB approval and research app registration. These safeguards help ensure that data collection can only occur with participant consent and should substantially reduce potential for misuse.

In line with best practices to safeguard participant privacy, researchers using the Stanford Screenomics platform should also adopt the principle of data minimization—collecting only the data that is strictly necessary for the research (Dunseath et al., 2018; Maeckelberghe et al., 2023; Nebeker et al., 2019, 2021). This principle should also extend to limiting data retention times and ensuring that data is only stored as long as required for legitimate research purposes, thus preventing the accumulation of unnecessary data that could later be misused (Nebeker et al., 2021). For the purposes of data sharing and management, summaries of key data features could be extracted and archived in place of raw data to support open science and reproducibility. Additionally, researchers should prioritize transparency with participants and consider conducting regular audits to ensure data are being used responsibly (Filkins et al., 2016; Nebeker et al., 2020). This includes being transparent about how data are analyzed within analytical algorithms to inform decisions. Algorithmic transparency helps mitigate concerns about opaque or biased decision-making processes and provides users with a clearer understanding of how their data are being used for research (Watson & Nations, 2019; Wrzus & Schoedel, 2023). By adhering to such rigorous ethical standards and maintaining oversight, the research community can help prevent harmful exploitation and ensure that the insights gained from the platform are used responsibly and for the public good.

## CONCLUSION

Using smartphone-based digital trace data offers a promising path toward providing more detailed insights into human behavior and well-being, reducing costs, understanding how smartphones are important in a broad spectrum of life, advancing personalized healthcare, and creating new theories of human behavior that require granular visibility into the details of moment-by-moment interactions. The Stanford Screenomics platform represents a significant step forward by offering an open-source, customizable framework for unobtrusive, multimodal data collection, including screenshots. By addressing limitations in scalability, customizability, and transparency found in existing tools—and by simultaneously capturing quantity-based and content-based data—it empowers researchers to conduct large-scale, flexible, cost-effective, and content-aware studies, yielding unprecedented insights into human behavior and health. With features like easy deployment, real-time monitoring, and adaptability to various research needs, the platform is poised for broad application across diverse research areas. As digital trace research continues to grow, we are committed to expanding the platform’s capabilities along with an open-source community, supporting collaboration, and ensuring reproducibility, ultimately enabling more comprehensive studies and facilitating the development of personalized interventions that improve health outcomes for individuals and populations.

## DECLARATIONS

### Ethics approval

Not applicable.

### Consent to participate

Not applicable.

### Consent for publication

All authors have reviewed the final manuscript and consent to its publication.

### Availability of data and materials

The text-based, illustrative 24-hour digital trace data collected using the Stanford Screenomics platform is available from the corresponding author upon request. The original screenshots are not available, as they have not been processed to protect the privacy of individuals whose information was recorded.

### Code availability

The source code for the Stanford Screenomics platform developed by I.K. and J.B. is available for public access at: https://github.com/iansulin/stanford_screenomics

### Authors’ contribution

I.K. conceived and designed the work, conducted data acquisition and analysis, interpreted data, drafted the manuscript, and performed critical revisions. I.K. and J.B. developed the software, with N.K. assisting in its testing. M.C. contributed to the conception and design of the work. D.E.C. contributed to data interpretation and critical revisions. N.H., T.N.R., B.R., and N.R. contributed to the conception and design, data interpretation, critical revisions, and secured funding. All authors reviewed and approved the final manuscript.

## Data Availability

The text-based, illustrative 24-hour digital trace data collected using the Stanford Screenomics platform is available from the corresponding author upon request. The original screenshot are not available, as they have not been processed to protect the privacy of individuals whose information was captured.

